# Development and Psychometric Validation of the Pelvic Dystonia Severity Scale (PDSS)

**DOI:** 10.64898/2026.07.24.26358500

**Authors:** Jason Siefferman, Mariia Safroshkina, Iuliia Nazarova, Andrey Zaznaev, Sameeha Hasan

**Affiliations:** Pain Management, Manhattan Pain Medicine, PLLC, New York, NY; Department of Biomedical Informatics, Columbia University, New York, NY

**Keywords:** Pelvic pain, pelvic floor disorders, patient reported outcome measure, psychometrics, focal dystonia, botulinum toxin

## Abstract

**Background:** Chronic pelvic pain with involuntary pelvic-floor hypertonicity is common, disabling, and inconsistently measured, and no validated condition-specific severity instrument exists. A refractory subset has been proposed to represent a focal dystonia of the pelvic musculature, termed *pelvic dystonia*.

**Purpose:** To develop the Pelvic Dystonia Severity Scale (PDSS) and evaluate its measurement properties as a patient-reported measure of pelvic-pain symptom severity and burden, following the COSMIN guidelines.

**Methods:** Cross-sectional study with a test–retest component in 102 adults from an outpatient multidisciplinary pain practice. We assessed data quality, structural validity, internal consistency, test–retest reliability, measurement error, and construct validity against the Global Dystonia Severity Rating Scale (GDS) and Brief Pain Inventory (BPI). Because no validated diagnostic criteria exist, a clinician blinded to PDSS responses rated each participant for clinical signs of pelvic dystonia (present/possible/absent) as a provisional reference standard.

**Results:** 100 of 102 participants (98%) returned complete data, with 0% item-level missing data. Factor analysis supported a unidimensional structure (single factor, 68.6% of variance; loadings 0.56–0.93), with high subscale intercorrelations (r = 0.81–0.97). Internal consistency (Cronbach’s α 0.810–0.924) and test–retest reliability (ICC 0.857–0.953) were strong. Convergent validity was supported by correlations with GDS pelvic-region items (r = 0.56–0.68) and BPI severity (r = 0.44–0.55), and discriminant validity by weak correlations with anatomically remote regions (shoulder/arm r = 0.03–0.16). PDSS scores rose monotonically across blinded clinical-signs categories (absent 14.9, possible 32.7, present 52.0; Kruskal–Wallis p < 0.001), discriminating signs-present from signs-absent participants with a very large effect (Hedges g = 2.19; ROC AUC = 0.92).

**Conclusion:** The PDSS is a psychometrically robust, unidimensional measure of pelvic-pain symptom severity and burden with strong data quality, reliability, and construct validity. It is suitable for characterizing symptom severity and, pending responsiveness testing, for monitoring treatment. The dystonia interpretation of the underlying phenotype is discussed as a hypothesis for future neurophysiologic and longitudinal study.

## Introduction

Dystonia is defined by sustained or intermittent muscle contractions causing abnormal, often repetitive movements, postures, or both, with an overall adult-onset prevalence of about 16 per 100,000 and comprising roughly 20% of movement-disorder clinic populations.^1–4^ Chronic pelvic pain with involuntary pelvic-floor hypertonicity is likewise common and frequently refractory, yet it is measured inconsistently across neurology, urology, pain management, and gynecology. It has been proposed that a subset of this phenotype — characterized by increased involuntary tone, pain, and impaired function that does not respond to conventional myofascial treatment — may represent a focal dystonia of the pelvic musculature.^5,6^

Symptom overlap with pelvic-pain and pelvic-floor dysfunction syndromes contributes to delayed recognition and inconsistent terminology, likely underestimating true prevalence.^7^ Whatever the ultimate nosology, the field lacks a standardized, quantitative, condition-specific measure of symptom severity, which limits phenotyping, outcome assessment, and research. Validated patient-reported outcome measures have advanced the study of other focal dystonias^8^; an analogous instrument for the hypertonic pelvic-pain phenotype is needed.

### Clinical Description

The phenotype of interest presents as involuntary pelvic-floor muscle hypertonicity, pain, restricted range of motion, and impaired function, including pelvic-floor tightness, dyspareunia, and urinary or bowel dysfunction, with context-dependent exacerbation during standing, ambulation, urination, defecation, or intercourse.^5,6,9^ A distinguishing clinical observation is that, in a subset of patients, the increased involuntary tone, pain, and functional impairment do not resolve with physical therapy or trigger-point injection and may improve with botulinum neurotoxin (BoNT); this treatment-response pattern has been interpreted as consistent with a dystonic mechanism.^9,10^ Placeholder diagnoses such as levator ani syndrome yield unreliable treatment outcomes. Recognizing PDys as a distinct pathophysiologic entity enables quantification and systematic study of treatment effects.

Framed within standard dystonia phenomenology — intermittent or sustained involuntary contractions producing abnormal postures and hypertonicity that fluctuate with activation, posture, stress, and attention^6^ — the proposed construct, like cervical and task-specific limb dystonia, would be expected to worsen with voluntary actions (standing, gait, toileting, intercourse) and to display patterned co-contraction rather than random spasm.^5^ Comprehensive examination includes palpation of the perineum, pelvic-floor muscles (obturator internus, levator ani, coccygeus), lumbosacral spine, sacrum, coccyx, and hip and sacroiliac joints. Physical signs proposed to parallel cervical and limb dystonia are summarized in **Table 1** and described below.

**Table 1.** Shared clinical features of pelvic, cervical, and limb dystonia.

| Physical sign | Description |
| --- | --- |
| Posture | Persistent or episodic pelvic tilt/rotation and abnormal lumbar lordosis or flattening driven by patterned contraction; sustained hip adduction, internal/external rotation, or flexion patterns that recur predictably; sustained or intermittent pelvic-floor contraction (“holding,” “clenching”) that is patterned and reproducible in specific contexts. |
| Movements | Repetitive involuntary contractions during mechanical shifts (sitting, standing, walking, positional transitions); sensed or palpated contractions of deep internal and/or pelvic-floor and perineal musculature; movements that appear “tremor-like” or “spasm-like,” risking misclassification. |
| Gestes antagonistes (“tricks”) | A light touch/pressure cue, positional change, or specific pelvic/hip adjustment that transiently relieves pelvic “pulling/clenching.” Relief occurs during the geste or soon after its initiation and may last as long as the maneuver. Physical therapy is often temporarily helpful, with symptoms returning soon after each session. |
| Overflow | Abdominal-wall bracing, hip co-contraction, gluteal tightening, or proximal-thigh guarding; inappropriate recruitment spreading into adjacent trunk musculature during provocative maneuvers (walking, jogging, turning, backward or figure-eight walking, stairs). |
| Mirror phenomena | Posture/contraction elicited or amplified when performing contralateral or remote motor tasks (e.g., complex contralateral leg/foot tasks provoking ipsilateral pelvic/hip contraction). |

*Features are adapted from established descriptions of focal (cervical and limb) dystonia and are proposed, not established, for the pelvic region.*

### Physical examination features supporting pelvic dystonia

#### Posture

As in cervical dystonia, sustained abnormal posture is a defining feature, although the pelvic region may require especially careful examination. It may present with asymmetric sacroiliac positioning, pelvic tilt, or hip rotation; contraction is often continuous, forcing the pelvis into a sustained posture that may change with repositioning.^5^

#### Movements

Abnormal movements include sustained or intermittent involuntary contractions of the pelvic musculature. The examiner should assess for involuntary pelvic-floor contraction elicited or amplified during sit-to-stand transfer, ambulation, or stair climbing, and for failure to voluntarily relax the pelvic floor between maneuvers. Across focal dystonias, abnormal posturing may worsen with voluntary action and produce compensatory maneuvers.^5^

#### Gestes antagonistes (“tricks”)

These are voluntary maneuvers that reduce abnormal contraction or posture. The examiner should document whether a light touch or pressure cue, positional change, or specific pelvic or hip adjustment transiently relieves pelvic “pulling” or “clenching.” Relief occurs during the geste or soon after its initiation and may last as long as the maneuver is maintained. Physical therapy is often temporarily helpful, although symptoms may return soon after each session.

#### Overflow

Overflow refers to unintentional contraction of neighboring regions accompanying the primary action - for example, abdominal-wall bracing, hip co-contraction, gluteal tightening, or proximal-thigh guarding during provocative maneuvers such as walking, jogging, turning, backward or figure-eight walking, or stair climbing. Recognizing overflow helps distinguish patterned contraction from volitional pain-guarding.

#### Mirror phenomena

Abnormal posture patterns (e.g., pelvic tilt, hip rotation) or contractions may be elicited or amplified by contralateral or remote motor tasks; the examiner may palpate reproducible ipsilateral pelvic or hip contraction that the patient does not voluntarily initiate.

Physical examination should document hypertonicity or spasm, trigger-point tenderness versus patterned contraction, asymmetry, and whether contraction appears task-linked (e.g., during voiding or defecation) rather than reflecting pain-guarding. In current terminology, the International Continence Society and IUGA describe increased pelvic-floor tone using the vocabulary of overactivity and hypertonicity rather than “dystonia”^7^; adoption of a dystonia framework for the pelvic floor is not yet established and is advanced here as a hypothesis rather than accepted terminology.

Proposed supportive criteria for the phenotype include: symptoms persisting >3 months; pain, involuntary hypertonicity, and functional impairment (bowel, bladder, sexual, mechanical); failure of ≥6 weeks of pelvic-floor physical therapy to resolve hypertonicity; inadequate or only temporary relief with oral muscle relaxants (e.g., baclofen, methocarbamol, diazepam, tizanidine); patterned contraction on examination; failure of trigger-point injection; and improvement with BoNT injection. We note that including BoNT response among these criteria introduces circularity when the same construct is later used to interpret treatment response, and we return to this in the Discussion.

### Pathophysiology of pelvic dystonia

Pelvic dystonia represents a focal dystonia and its mechanism is expected to parallel those of dystonia broadly, which may be primary (idiopathic) or secondary (acquired). Proposed contributors include genetic susceptibility (variants in TOR1A, THAP1, and related genes affecting neurodevelopmental pathways and striatal dopaminergic signaling), altered sensory processing, trauma, and impaired basal-ganglia function.^1,11^ Acquired triggers described for secondary dystonia include complex regional pain syndrome, trauma, chronic guarding, childbirth, surgery, pelvic inflammatory conditions, autoimmune disease, infection, vascular insults, and drugs or toxins.^12^ Altered GABAergic, dopaminergic, and cholinergic signaling implicated in dystonia generally may be relevant, although the pathogenesis of any pelvic presentation remains incompletely understood.^1,6^

Secondary presentations may become persistent through maladaptive neuroplasticity, progressively less responsive to myofascial or pain-oriented approaches (physical therapy, acupuncture, trigger-point injection, muscle relaxants).^3,6,9^ When dystonia is the primary driver, conventional symptomatic treatment may provide incomplete relief because it does not address abnormal movement and posture, and focal-dystonia guidelines accordingly emphasize targeted therapies.^2,3^ Importantly, not all pelvic pain with hypertonicity is dystonic: BoNT outcomes are variable in conditions such as levator ani syndrome and high-tone pelvic-floor dysfunction,^10,13,14^ underscoring the need for careful phenotyping before attributing refractory pelvic pain to a dystonic mechanism and the need for a quantitative severity measure to support such phenotyping.

### Statement of Purpose

This study develops and validates the PDSS, a condition-specific instrument designed to quantify symptom severity and functional burden in the hypertonic pelvic-pain phenotype and to monitor treatment outcomes. Because this phenotype overlaps symptomatically with other pelvic-pain conditions and no validated diagnostic reference standard exists, we deliberately separate measurement from diagnosis. A validated PDSS will: (1) provide a standardized, quantitative measure of symptom severity and burden; (2) enable objective assessment of treatment response (e.g., to physical therapy or BoNT); and (3) support research to refine phenotyping and, ultimately, to test whether a dystonic subtype can be defined.

**Study questions:** (1) Can the PDSS reliably quantify symptom severity and functional impact? (2) Does the PDSS demonstrate sound measurement properties — data quality, structural validity, internal consistency, temporal stability, and construct validity — relative to the GDS and BPI? (3) Is the PDSS best represented by a single severity dimension or by separable Pain, Muscle Tone, and Functional Impairment subscales?

## Methods

### Setting and Participants

This observational psychometric validation study used a cross-sectional design with a test–retest (TRT) component. Participants were recruited from an outpatient multidisciplinary pain practice experienced in treating pelvic pain of all sources, including the suspected hypertonic/dystonic phenotype.

**Inclusion criteria:** adults ≥18 years of any sex, with or without current pelvic-floor dysfunction symptoms.

**Exclusion criteria:** non-English speakers; impaired mental capacity; current pregnancy or delivery within 6 months; abdominal or pelvic surgery within 6 months; or a neuromuscular or movement-disorder diagnosis other than dystonia (e.g., myasthenia, muscular dystrophy, myopathy).

### Participant classification and reference standard

Pelvic dystonia is not currently included in consensus classifications of dystonia^5,11^ and has no validated diagnostic criteria; consequently, no reference standard exists against which participants could be classified as confirmed cases, and we do not report a proportion “diagnosed,” as doing so would presuppose the nosological status that remains to be established. Instead, to characterize the sample and to permit known-groups analysis, each participant was classified by clinical symptom status as symptomatic (presenting with chronic pelvic pain and examination evidence of pelvic-floor hypertonicity) or as a comparison participant (without current pelvic-floor symptoms).

To characterize the sample and to permit known-groups analysis, a clinician rated each participant for the presence of clinical signs of pelvic dystonia on examination, using a three-level provisional classification — signs present (“yes”), equivocal (“possible”), or absent (“no”). The rating clinician was blinded to participants’ PDSS responses, so that this clinician-assessed classification served as a reference standard independent of the patient-reported PDSS.

### Measurements

The PDSS is a disease-specific 10-item instrument organized into three content areas — Pain (3 items), Muscle Tone (3 items), and Functional Impairment (4 items; partially adapted from the Oswestry Disability Index)^15^. Each item uses a 10-point Likert scale (0 = no symptom; 10 = worst imaginable)^16^; total and content-area scores are unweighted sums, with higher scores indicating greater severity. Candidate items were developed by several physicians and were grounded in an extensive review of the focal-dystonia and pelvic-pain literature, in systematic chart review to catalog recurring presenting symptoms, in adaptation from existing validated instruments (the Functional Impairment items were adapted from the Oswestry Disability Index^15^), and in collective clinical experience, so as to span the pain, involuntary-tone, and functional dimensions of the phenotype. Formal content-validity procedures — structured expert-panel rating of item relevance and comprehensiveness (e.g., a content-validity index or Delphi process) and patient cognitive interviewing to confirm comprehensibility and comprehensiveness — were not conducted in this initial phase and are planned as the next development step.^17^

The psychometric evaluation was designed within the consensus-based standards for the selection of health measurement instruments (COSMIN) framework for patient-reported outcome measures.^17,18^ Consistent with COSMIN guidance for an initial validation, and to keep the report focused, we prioritized structural validity, internal consistency, test–retest reliability, measurement error, and construct validity. Due to the cross-sectional study design and the fact that it was performed in a single English-speaking clinical site, responsiveness and cross-cultural validity were reserved for future investigation.

Convergent and discriminant validity were assessed against the GDS, which rates dystonia severity across 10 body regions on 0–10 scales (maximum 140),^19^ and the short-form BPI, which assesses pain severity and interference with daily activities on 0–10 scales.^20^

### Data Analysis

Five psychometric properties were examined: data quality, scaling assumptions, targeting, reliability, and validity.^21–23^ Data quality was assessed through response completeness and missing-data patterns. Scaling assumptions were evaluated with corrected item–total and item– other-scale correlations to determine whether items could be validly aggregated. Targeting was assessed via score distributions and floor/ceiling effects. Reliability was evaluated through internal consistency (Cronbach’s α^24^) and test–retest reproducibility (ICC₂,₁, absolute agreement^25^), with measurement error summarized by the standard error of measurement and minimal detectable change; precision of the reliability estimate was interpreted using confidence intervals rather than post-hoc power.^26^ Structural validity was examined by exploratory factor analysis with parallel analysis.

Construct validity was evaluated by testing pre-specified hypotheses about the direction and magnitude of correlations between the PDSS and comparator measures (Table 5), reporting the proportion confirmed against the COSMIN ≥75% criterion. We hypothesized moderate-to-strong correlations with the GDS pelvis/proximal-leg region and with BPI severity (convergent), and weak correlations with anatomically remote GDS regions (discriminant). Known-groups (discriminant) validity was assessed by comparing PDSS total scores across the clinician-assessed clinical-signs classification (present/possible/absent) using the Kruskal–Wallis test and an ordered-trend test (Spearman correlation of PDSS score with the ordinal classification), with pairwise effect sizes (Hedges g) and the area under the ROC curve for discriminating signs-present from signs-absent participants. Analyses were performed in Python using the pandas,^27^ NumPy,^28^ and pingouin^29^ libraries.

## Results

### Sample, composition, and data quality

Of 102 participants who received the PDSS, 100 returned complete questionnaires (corrected response rate 98%); all 44 TRT questionnaires were returned (100%). By clinician-assessed clinical signs of pelvic dystonia, 39 participants (39%) were classified as signs present, 29 (29%) as possible, and 32 (32%) as signs absent; 2 participants were unclassified and were excluded from known-groups analyses. Item-level missing data were 0% across all PDSS items, and total and content-area scores were computable for all 100 participants, indicating excellent data quality (**Supplementary Table 1**). TRT participants completed the PDSS a second time approximately one week after the initial assessment.

### Scaling assumptions and structural validity

Corrected item–total correlations ranged from 0.476 to 0.879 (all Spearman ρ > 0.30; **Supplementary Table 1**). However, items correlated with competing content areas approximately as strongly as with their own (mean item–own vs item–other: Pain 0.83 vs 0.82; Muscle Tone 0.85 vs 0.84; Functional Impairment 0.64 vs 0.65), indicating that the three content areas do not function as statistically separable subscales. Consistent with this, exploratory factor analysis showed excellent sampling adequacy (Kaiser–Meyer–Olkin = 0.891) and a significant Bartlett’s test (χ² = 1187, df = 45, p < 0.001), and parallel analysis and the scree plot (**Figure 1**) supported a single factor: only the first eigenvalue exceeded 1 (6.86), explaining 68.6% of variance, with all items loading strongly (0.56–0.93; highest for Pain and Muscle Tone items, 0.88–0.93; **Supplementary Table 3**). Subscale intercorrelations were correspondingly high (r = 0.81–0.97; Pain–Muscle Tone r = 0.97). We therefore treat the PDSS as a unidimensional severity measure summarized by a single total score, retaining Pain, Muscle Tone, and Functional Impairment as descriptive content facets rather than independently scored dimensions.

**Figure 1.**
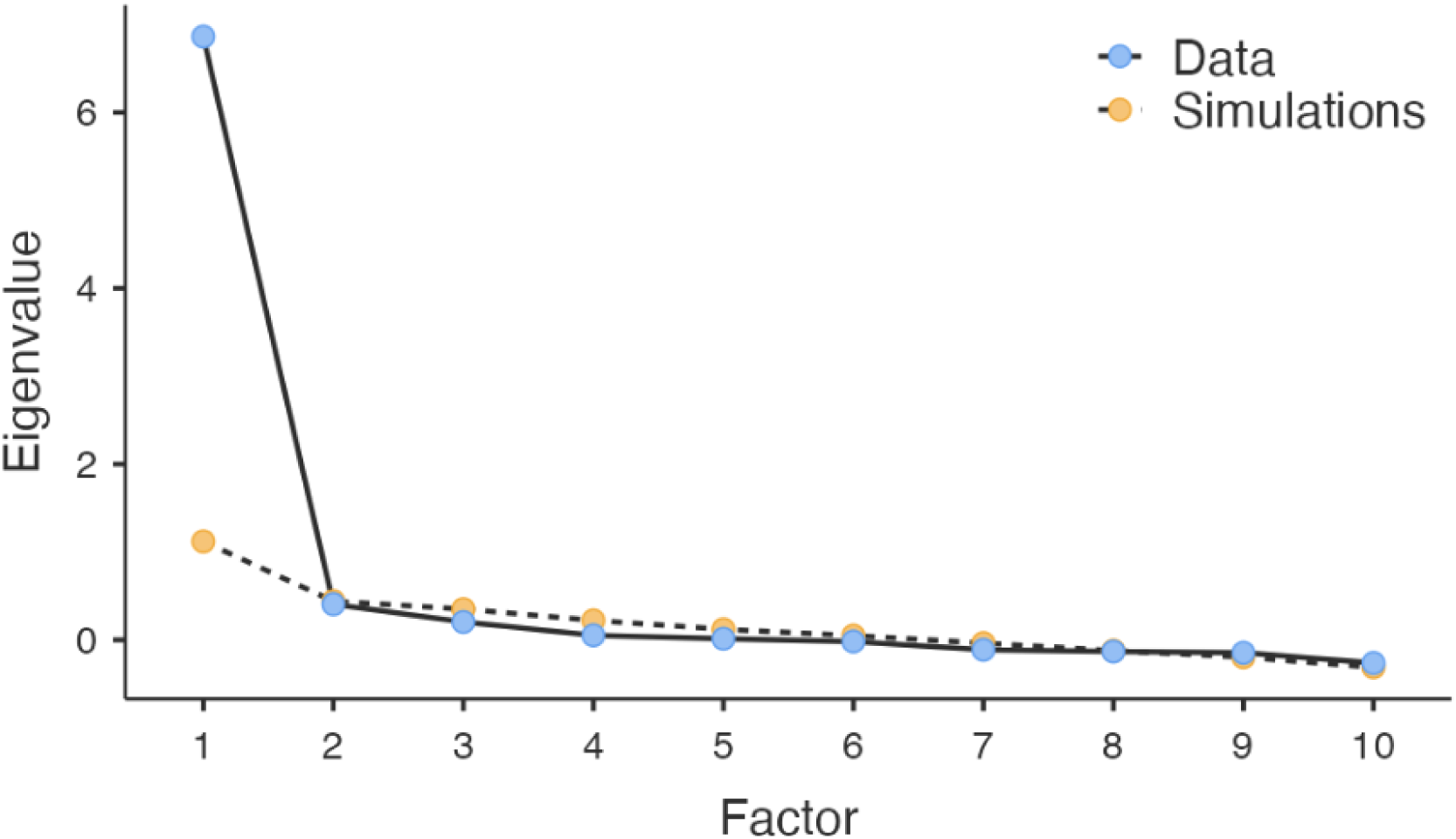
Scree plot from exploratory factor analysis. Only the first eigenvalue exceeds 1, supporting a unidimensional structure of the PDSS.

### Targeting

The Pain and Muscle Tone facets showed full score-range utilization (0–30), low floor effects (9.8% and 11.8%), minimal ceiling effects (0.98%), and near-symmetric distributions (skewness −0.096 and −0.033; kurtosis −1.19 and −1.18). The Functional Impairment facet reached a maximum of 34/40 and showed a floor effect of 24.5% (exceeding the 15% criterion), indicating that many participants reported minimal functional impairment; ceiling effect (0.98%), skewness (0.591), and kurtosis (−0.617) remained acceptable. The floor effect is consistent with the enrollment of participants with and without current symptoms and should be re-examined within the symptomatic subgroup.

### Reliability and measurement error

Internal consistency was high (Cronbach’s α: Pain 0.912, Muscle Tone 0.924, Functional Impairment 0.810), interpreted in light of the unidimensional structure established above. Test– retest reliability over approximately one week was strong (ICC₂: Pain 0.878 [95% CI 0.78–0.93], Muscle Tone 0.857 [0.74–0.92], Functional Impairment 0.953 [0.91–0.98]). Measurement error was low relative to scale range (SEM 2.83–3.18; MDC₉₅ 6.28–8.81), supporting sensitivity to meaningful change. The 95% confidence intervals for the ICCs, based on 44 test–retest participants, exclude values below the 0.70 threshold and thus support adequate reliability.^26^

### Construct validity: convergent and discriminant

Pre-specified hypotheses and results are shown in **Supplentary Table 4**. Convergent validity was supported: the PDSS correlated r = 0.56–0.68 with GDS pelvis/proximal-leg items (Pain– right 0.63, Pain–left 0.68; Muscle Tone–right 0.63, Muscle Tone–left 0.67) and r = 0.44–0.55 with BPI severity items (**Supplementary Table 2**), consistent with expectations for a pelvic-specific severity measure. Discriminant validity was supported at the extremes: correlations with GDS shoulder/proximal-arm items were weak (r = 0.03–0.16), and the pelvic region correlated more strongly than remote regions. Correlations with intermediate, anatomically unrelated regions (face, jaw, larynx, trunk) were low-to-moderate (r = 0.30–0.55), and the Functional Impairment facet correlated 0.55 with GDS eyes/upper-face; we interpret this residual cross-regional association as a general dystonia-severity or symptom-reporting tendency in the GDS rather than pelvic-specific overlap. Overall, 5 of 6 pre-specified hypotheses were met (83%), above the COSMIN ≥75% threshold.

### Known-groups (discriminant) validity

PDSS total scores increased monotonically across the clinician-assessed classification (signs absent: mean 14.9 [SD 16.7], median 10; possible: 32.7 [22.4], median 36; signs present: 52.0 [16.9], median 55; Table 6). The three groups differed significantly (Kruskal–Wallis H = 42.9, p < 0.001), with a strong ordered trend (Spearman ρ = 0.66, p < 0.001). The PDSS discriminated signs-present from signs-absent participants with a very large effect (Hedges g = 2.19; Mann– Whitney p < 0.001) and excellent accuracy (ROC AUC = 0.92); adjacent contrasts were also large (present vs possible g = 0.99; possible vs absent g = 0.89). The intermediate position of the “possible” group — in whom clinical signs are equivocal and a diagnostic botulinum-toxin injection would help distinguish true pelvic dystonia from other causes of hypertonic pelvic pain — is consistent with the PDSS tracking a graded underlying construct. Some score overlap between groups is expected of a severity measure and is consistent with the PDSS quantifying symptom burden rather than functioning as a stand-alone diagnostic test.

## Discussion

This study provides psychometric validation of the PDSS as a reliable, unidimensional measure of pelvic-pain symptom severity and burden. The instrument showed excellent data quality, high internal consistency, strong test–retest reliability, low measurement error, and construct validity supported by pre-specified hypotheses.^21–23^ Rather than three separable domains, the data showed a single-factor solution explaining 68.6% of variance, high subscale intercorrelations (r = 0.81–0.97), and item–own correlations comparable to item–other correlations — indicate a single severity construct; we therefore recommend a total score, with Pain, Muscle Tone, and Functional Impairment retained as descriptive facets. The Functional Impairment facet was more heterogeneous and showed a floor effect (24.5%), yet retained acceptable distributional properties and clinical informativeness, particularly at moderate-to-severe symptom burden.

Convergent validity was strongest where clinically expected — with the GDS pelvis/proximal-leg region and with BPI severity — while correlations with anatomically remote regions were weak, supporting discriminant validity. The residual low-to-moderate correlations with unrelated regions most plausibly reflect a general severity or symptom-reporting tendency captured by the GDS, which rates dystonia across the whole body; a pelvic- or pain-specific comparator, or a measure expected to be uncorrelated with pelvic severity, would sharpen future discriminant testing. Prior work has emphasized persistent difficulty distinguishing this phenotype from pelvic-floor dysfunction, chronic pelvic pain, and myofascial disorders^7,14^; a standardized, empirically supported severity measure is a step toward more consistent phenotyping and outcome assessment.

Several limitations warrant emphasis. First, and most importantly, pelvic dystonia is not an established diagnosis: it is absent from consensus dystonia classifications,^5,11^ and current pelvic-floor terminology describes increased tone as hypertonicity or overactivity rather than dystonia.^7^ Accordingly, we frame the PDSS as a severity measure for a clinically observable hypertonic pelvic-pain phenotype and treat the dystonia interpretation as a hypothesis requiring neurophysiologic confirmation (e.g., patterned electromyographic co-contraction) and expert consensus. Second, pelvic dystonia has no validated diagnostic criteria; the clinician-assessed clinical-signs classification used here is a provisional reference standard, and the known-groups evidence is conditional on it. Reassuringly, the rating clinician was blinded to PDSS responses, so the group separation cannot be attributed to shared-source bias; nonetheless, independent replication with multiple raters and reported inter-rater agreement would further strengthen this evidence. Third, several proposed supportive criteria include BoNT response, which introduces circularity with the treatment the scale may later be used to monitor; future work should test the construct against treatment-independent criteria. Fourth, the cohort was drawn from a single clinical site, limiting generalizability. Fifth, responsiveness was not evaluated; longitudinal studies using distribution- and anchor-based methods (e.g., standardized response mean, minimal important change) are needed before the PDSS is used to quantify treatment effects.^21–23^ Sixth, although item development drew on literature review, chart review, established instruments, and multi-clinician experience, formal content-validity assessment — structured expert consensus and patient cognitive interviewing — was not undertaken in this phase; because content validity is the most important measurement property, establishing it is a priority next step.^17^

Despite these limitations, the study delivers a validated severity measure for a common and difficult-to-quantify problem and provides preliminary, hypothesis-generating support for the possibility that a subset of refractory hypertonic pelvic pain reflects a focal dystonia. A standardized instrument enables the multi-site, longitudinal, and neurophysiologic studies required to test that hypothesis rigorously.

## Conclusion

We introduce the PDSS as a standardized, unidimensional patient-reported measure of pelvic-pain symptom severity and burden. Across multiple analyses it demonstrated strong reliability and construct validity, supporting its use for symptom characterization and, pending responsiveness testing, treatment monitoring. By quantifying a phenotype that is currently measured inconsistently, the PDSS can improve clinical assessment and provide the common metric needed for future research — including studies designed to test whether a dystonic subtype of pelvic-floor hypertonicity can be defined and validated.^14,22,23^

## Ethics Approval and Informed Consent

The study protocol was approved by the Biomedical Research Alliance of New York (BRANY) institutional review board. All participants provided informed consent before enrollment, either by digital signature (HIPAA-compliant Jotform platform) or by signing a paper form.

## Data Availability

The datasets generated and analyzed during the current study are available from the corresponding author on reasonable request.

## Author Contributions

*Conceptualization*: JS. *Methodology*: JS, MS. *Data collection*: MS, IN. *Formal analysis and validation*: AZ, MS, IN. *Writing* (original draft): JS, MS. *Writing* (review & editing): JS, MS, IN, AZ, SH. *Visualization*: MS, IN, AZ, SH. *Supervision*: JS, MS. *Project administration*: MS

## Acknowledgements

The authors thank the study participants, and the clinical and administrative staff at Manhattan Pain Medicine, PLLC, for their support with study implementation, participant coordination, and data collection. All individuals acknowledged have provided permission to be named.

## Supplementary Tables

**Supplementary Table 1.** Targeting, reliability, and measurement error of the PDSS content facets.

| Psychometric property | Pain (3 items) | Muscle Tone (3 items) | Functional Impairment (4 items) |
| --- | --- | --- | --- |
| Data quality — item missing data (%) / computable scores (%) | 0% / 100% | 0% / 100% | 0% / 100% |
| Corrected item–total correlations — mean (range) | 0.834 (0.818–0.869) | 0.854 (0.833–0.879) | 0.644 (0.476–0.718) |
| Item–other-scale correlations — mean (range) | 0.823 (0.719–0.921) | 0.838 (0.747–0.936) | 0.654 (0.526–0.778) |
| Targeting — mean (SD) | 12.1 (8.05) | 12.0 (8.23) | 10.2 (9.01) |
| Score range / floor % / ceiling % | 0–30 / 9.80 / 0.98 | 0–30 / 11.8 / 0.98 | 0–34 / 24.5 / 0.98 |
| Skewness / kurtosis | –0.096 / –1.19 | –0.033 / –1.18 | 0.591 / –0.617 |
| Cronbach’s $\alpha$ | 0.912 | 0.924 | 0.810 |
| Test–retest ICC<sub>2</sub> (95% CI) | 0.878 (0.78–0.93) | 0.857 (0.74–0.92) | 0.953 (0.91–0.98) |

*SD = standard deviation; ICC = intraclass correlation coefficient; CI = confidence interval. Content facets are reported descriptively; the validated score is the unidimensional PDSS total.*

**Supplementary Table 2.** Convergent and discriminant validity: correlations of PDSS facets with the GDS and BPI.

| **Instrument / region** | **Pain facet** | **Muscle Tone facet** | **Functional Impairment facet** |
| --- | --- | --- | --- |
| PDSS — Pain facet | 1 | 0.97 | 0.81 |
| PDSS — Muscle Tone facet | 0.97 | 1 | 0.82 |
| PDSS — Functional Impairment facet | 0.81 | 0.82 | 1 |
| GDS — Eyes and upper face | 0.34 | 0.38 | 0.55 |
| GDS — Lower face | 0.32 | 0.37 | 0.47 |
| GDS — Jaw and tongue | 0.30 | 0.36 | 0.37 |
| GDS — Larynx | 0.32 | 0.36 | 0.33 |
| GDS — Neck | 0.14 | 0.21 | 0.27 |
| GDS — Shoulder/proximal arm (R) | 0.03 | 0.11 | 0.09 |
| GDS — Shoulder/proximal arm (L) | 0.11 | 0.16 | 0.16 |
| GDS — Distal arm/hand (R) | 0.25 | 0.29 | 0.31 |
| GDS — Distal arm/hand (L) | 0.23 | 0.25 | 0.28 |
| GDS — Pelvis/proximal leg (R) | 0.63 | 0.63 | 0.60 |
| GDS — Pelvis/proximal leg (L) | 0.68 | 0.67 | 0.56 |
| GDS — Distal leg/foot (R) | 0.39 | 0.39 | 0.41 |
| GDS — Distal leg/foot (L) | 0.32 | 0.33 | 0.32 |
| GDS — Trunk | 0.46 | 0.50 | 0.50 |
| BPI — Severity Q3 | 0.44 | 0.43 | 0.44 |
| BPI — Severity Q4 | 0.40 | 0.38 | 0.32 |
| BPI — Severity Q5 | 0.55 | 0.53 | 0.50 |
| BPI — Severity Q6 | 0.50 | 0.48 | 0.51 |
| BPI — Interference Q9A | 0.53 | 0.51 | 0.57 |
| BPI — Interference Q9B | 0.45 | 0.42 | 0.41 |
| BPI — Interference Q9C | 0.34 | 0.34 | 0.38 |
| BPI — Interference Q9D | 0.36 | 0.37 | 0.47 |
| BPI — Interference Q9E | 0.50 | 0.48 | 0.53 |
| BPI — Interference Q9F | 0.36 | 0.35 | 0.43 |

| Instrument / region | Pain facet | Muscle Tone facet | Functional Impairment facet |
| --- | --- | --- | --- |
| BPI — Interference Q9G | 0.50 | 0.47 | 0.46 |

*Values are correlation coefficients. Shaded rows indicate the a priori convergent target (GDS* *pelvis/proximal leg). GDS = Global Dystonia Severity Rating Scale; BPI = Brief Pain Inventory;* *R/L = right/left.*

**Supplementary Table 3.** Exploratory factor analysis of PDSS items: loadings, uniqueness, and sampling adequacy.

| Item | Factor loading | Uniqueness | KMO (item) |
| --- | --- | --- | --- |
| Pain — frequency | 0.881 | 0.224 | 0.926 |
| Pain — at rest | 0.901 | 0.189 | 0.849 |
| Pain — during activity | 0.896 | 0.198 | 0.858 |
| Tone — frequency | 0.878 | 0.229 | 0.927 |
| Tone — at rest | 0.927 | 0.142 | 0.853 |
| Tone — during activity | 0.922 | 0.151 | 0.859 |
| Function — sitting | 0.697 | 0.514 | 0.904 |
| Function — sex | 0.820 | 0.328 | 0.965 |
| Function — travel | 0.724 | 0.476 | 0.901 |
| Function — bathroom | 0.560 | 0.686 | 0.924 |

*A single factor was extracted (68.6% of variance). KMO = Kaiser–Meyer–Olkin measure of sampling adequacy (overall KMO = 0.891).*

**Supplementary Table 4.** Pre-specified construct-validity hypotheses and results (COSMIN).

| # | Hypothesis (PDSS vs ...) | Predicted | Observed | Met? |
| --- | --- | --- | --- | --- |
| H1 | GDS pelvis/proximal-leg items (target region) | $r \geq 0.50$ | 0.56–0.68 | **Yes** |
| H2 | GDS shoulder/proximal-arm items (remote region) | $r < 0.30$ | 0.03–0.16 | **Yes** |
| H3 | Stronger with pelvic than remote region | pelvic > arm | $\approx 0.65$ vs $\approx 0.10$ | **Yes** |
| H4 | BPI pain-severity items | $r \geq 0.40$ | 0.40–0.55 | **Yes** |
| H5 | BPI pain-interference items | moderate | 0.34–0.57 | **Yes** |
| H6 | GDS remote face/jaw/larynx (discriminant probe) | $r < 0.30$ | 0.30–0.55 | **No\*** |

*Observed values are taken across PDSS facets (Supplementary Table 2), recomputed against the* *PDSS total once scored.*
*\*H6 not met — interpreted as general dystonia-severity/method variance in the GDS, discussed* *as a discriminant limitation. Result: 5 of 6 (83%) met, above the COSMIN $\geq 75\%$ threshold.*

